## Supplementary for "Diagnostic testing preferences can help inform future public health response efforts: global insights from an international survey"

**Supplementary Table 1. Types of tests for the detection of SARS-CoV-2**[1]

| Type of Test | Description | Role in the Pandemic |
|---|---|---|
| Nucleic Acid Amplification Tests (NAATs) | Designed to detect at least one viral RNA gene, used to identify an infection that is current of recent by amplifying the RNA of the virus if any is present in an individual's sample | How diagnostic tests are able to identify individuals infected with SARS-CoV-2<br>Reliable, detect small amounts of SARS-CoV-2, unlikely to give a false positive result<br>High sensitivity and specificity |
| Antigen | Immunoassays that detect infection of SARS-CoV-2 by detecting the presence of a particular viral antigen | Have enabled at-home testing to become more widespread; rapid test results are beneficial for testing immediately before travel or gathering with others<br>Similar specificity to NAATs, but less sensitive<br>Less expensive than NAATs, may require follow up NAAT to confirm result |
| Antibody serology testing) | Determines if an individual has detectable antibodies against SARS-CoV-2 | A positive test indicates the presence of antibodies to SARS-CoV-2, which means it is possible an individual was recently infected and developed immunity to SARS-CoV-2<br>Help scientists gain an understanding of how the human immune system responds to SARS-CoV-2 and retrospectively assess the attack rate or size of an outbreak |

**Supplementary Table 2. Sample types commonly used for the detection of SARS-CoV-2[2–4] [supporting images available upon request from the authors].**

| Sample Type | Collection Method | Pros | Cons | Diagnostic Capability |
|---|---|---|---|---|
| Anterior-nares (AN) nasal swab | Swab is inserted into a patient's nose and rubbed in a circular motion with moderate pressure against the wall of the nares region. | Can be collected by a healthcare provider or self-collected by the patient. | Lower sensitivity compared to the nasopharyngeal swab. | 62.7-90.5% sensitivity[5] |
| Nasopharyngeal | Swab is inserted into a patient's nostril until it touches the nasopharynx, the upper part of the throat that lies behind the nose.<br>Swab is gently rotated and rubbed against the nasopharynx with the goal of absorbing respiratory secretions and infected cells if they are present. | High sensitivity | Requires trained healthcare professionals in full PPE to collect<br>Requires special swabs that can be affected by supply chain constraints, making testing more limited and challenging<br>Can induce gagging, coughing, or sneezing, potentially aerosolizing the virus. | 74.4-96.5% sensitivity[5]<br>(Reference sample type for US FDA, so trusted source for diagnosing respiratory viruses) |
| Oral (mouth swab) | Must refrain from food, drink or mouthwash for 20 minutes prior.<br>For 20-30 seconds, swab the inside of each cheek, the upper and lower gums, underneath and top of the tongue, back of the throat, and roof of the mouth.[6] | Can be collected by a healthcare provider or self-collected by the patient | | 98% sensitivity[7] |
| Oropharyngeal (throat swab) | Swab is inserted into a patient's mouth and rubbed over both tonsillar pillars and oropharynx. | | Requires trained healthcare professional in full PPE to collect.<br>Can induce coughing or gagging. | 70.3-94.7% sensitivity[5] |
| Saliva | Saliva is pooled in the mouth and drooled into a sample tube. | Can be self-collected by the patient. | | 92-98% sensitivity[8] |
| Serology test | An antibody or serology test is a blood test that looks for signs of a previous COVID-19 infection. It detects antibodies, which are proteins in the blood that fight off infection. Antibody testing has a lot of promise because it will help us understand the pervasiveness of COVID-19 in our communities.[9] | | Must be collected in a doctor's office or clinic, and requires a trained healthcare professional for collection. | Not generally useful for diagnosing COVID-19<br>96% sensitivity based on performance evaluations.[10] |

**Supplementary Table 3. Q9 - Q11 Keywords and Categories for Analysis**

| Question | Categories | Keywords |
|---|---|---|

| | | |
|---|---|---|
| Q9 | PCR | ‘pcr’ |
| | Lateral Flow Test | 'lateral', 'flow', lft' |
| | Saliva | ‘saliva’ |
| | Rapid Antigen Test | 'rapid', 'antigen', 'rat' |
| | Labcorp | 'labcorp' |
| | Multiple | |
| | Other | |
| Q10 | No preference | 'no', 'any' |
| | Abbott | ‘abbott', 'abbot', 'binax' |
| | Roche | 'roche' |
| | Labcorp | 'labcorp' |
| | Quidel | ‘quidel' |
| | Multiple | |
| | Other | |
| Q11 | Ease | 'ease', 'convenien', 'availabil','comfort', 'easy', 'access' |
| | Speed | 'speed', 'quick', 'fast', ‘time’ |
| | Accuracy | 'accur' |
| | Sensitivity | 'sensitivity', |
| | Reliability | 'reliable' |
| | Multiple | |
| | Other | |

**Supplementary Table 4. Geographical data for survey sample**

| Region | Responses (n) |
|---|---|
| **Asia** | **166 total** |
| Bangladesh | 3 |
| Bhutan | 0 |
| Cambodia | 2 |
| China | 8 |
| India | 35 |
| Indonesia | 28 |
| Japan | 6 |
| Kazakhstan | 3 |
| Malaysia | 18 |
| Myanmar | 3 |

| | |
|---|---|
| Nepal | 3 |
| Pakistan | 7 |
| Philippines | 7 |
| Singapore | 6 |
| South Korea | 5 |
| Sri Lanka | 4 |
| Taiwan | 11 |
| Tajikistan | 1 |
| Thailand | 12 |
| Vietnam | 4 |
| **Europe** | **409 total** |
| Austria | 1 |
| Belgium | 1 |
| Bosnia & Herzegovina | 17 |
| Bulgaria | 1 |
| Croatia | 1 |
| Czech Republic | 2 |
| Denmark | 3 |
| Estonia | 1 |
| Finland | 3 |
| France | 2 |
| Georgia | 3 |
| Germany | 20 |
| Greece | 9 |
| Hungary | 1 |
| Ireland | 8 |
| Italy | 21 |
| Latvia | 2 |
| Lithuania | 1 |
| Luxembourg | 0 |
| Netherlands | 9 |
| Norway | 2 |
| Poland | 8 |
| Portugal | 3 |
| Romania | 3 |
| Russia | 14 |
| Serbia | 0 |

| | |
|---|---|
| Slovakia | 1 |
| Slovenia | 1 |
| Spain | 25 |
| Sweden | 3 |
| Switzerland | 6 |
| United Kingdom of Great Britain and Northern Ireland | 237 |
| **Latin America & The Caribbean** | **186 total** |
| Argentina | 18 |
| Barbados | 1 |
| Bolivia | 1 |
| Brazil | 29 |
| Chile | 7 |
| Columbia | 12 |
| Costa Rica | 1 |
| Ecuador | 10 |
| El Salvador | 1 |
| French Guiana | 1 |
| Guatemala | 9 |
| Guyana | 2 |
| Honduras | 2 |
| Mexico | 67 |
| Paraguay | 11 |
| Peru | 8 |
| Saint Kitts and Nevis | 2 |
| Trinidad and Tobago | 1 |
| Uruguay | 2 |
| Venezuela | 1 |
| **North America** | **536 total** |
| Canada | 51 |
| United States | 485 |
| **Africa** | **449 total** |
| Benin | 1 |
| Botswana | 1 |
| Burkina Faso | 1 |
| Cameroon | 2 |
| Democratic Republic of the Congo | 4 |
| Ethiopia | 22 |

| | |
|---|---|
| Gabon | 1 |
| Ghana | 20 |
| Kenya | 219 |
| Lesotho | 1 |
| Liberia | 1 |
| Libya | 1 |
| Madagascar | 2 |
| Mali | 3 |
| Mauritius | 1 |
| Morocco | 4 |
| Mozambique | 2 |
| Nigeria | 93 |
| Rwanda | 1 |
| Senegal | 4 |
| Somalia | 1 |
| South Africa | 19 |
| South Sudan | 1 |
| Sudan | 5 |
| Tanzania | 1 |
| The Gambia | 1 |
| Tunisia | 4 |
| Uganda | 24 |
| Zambia | 6 |
| Zimbabwe | 3 |
| **Middle East** | **162 total** |
| Armenia | 3 |
| Azerbaijan | 0 |
| Bahrain | 3 |
| Egypt | 12 |
| Iran | 18 |
| Iraq | 4 |
| Israel | 4 |
| Jordan | 2 |
| Kuwait | 1 |
| Lebanon (Beirut) | 1 |
| Oman | 3 |
| Palestine | 58 |

| | |
|---|---|
| Qatar | 3 |
| Saudi Arabia | 18 |
| Syria | 2 |
| Turkey | 20 |
| United Arab Emirates | 7 |
| Yemen | 3 |
| **Oceania** | **120 total** |
| Australia | 19 |
| New Zealand | 99 |
| Papua New Guinea | 2 |

**Supplementary Table 5. Number of responses per question**

| Question | Number of responses received (n) |
|---|---|
| Question 1 | 2094 |
| Question 2 | 2094 |
| Question 3 | 2094 |
| Question 4 | 2094 |
| Question 5 | 2092 |
| Question 6 | 19 |
| Question 7 | 150 |
| Question 8 | 2094 |
| Question 9 | 2017 |
| Question 10 | 1903 |
| Question 11 | 2032 |
| Question 12 | 2094 |
| Question 13 | 1781 |
| Question 14 | 525 |
| Question 15 | 2094 |
| Question 16 | 2094 |
| Question 17 | 2080 |
| Question 18 | 2081 |
| Question 19 | 2094 |
| Question 20 | 2094 |
| Question 21 | 2021 |

**Supplementary Table 6. Most preferred diagnostic testing method in Asia**

| Diagnostic Testing Method | Mean Values Of Preference Rank* |
|---|---|
| Saliva | 2.81 |
| Anterior-nares (AN) nasal swab | 3.14 |
| Oropharyngeal | 3.57 |
| Oral swab | 3.08 |
| Nasopharyngeal | 3.79 |
| Deep coughing | 4.65 |
| **Top Preference by Country** | |
| Bangladesh | Nasopharyngeal |
| Bhutan | N/A |
| Cambodia | Oral Swab/Saliva |
| China | Saliva |
| India | AN swab |
| Indonesia | Nasopharyngeal |
| Japan | Saliva |
| Kazakhstan | Deep coughing |
| Malaysia | Saliva |
| Myanmar | Nasopharyngeal |
| Nepal | AN swab |
| Pakistan | AN swab |
| Philippines | Saliva |
| Singapore | Saliva |
| South Korea | Saliva |
| Sri Lanka | Saliva |
| Taiwan | Saliva |
| Tajikistan | AN swab |
| Thailand | Oral swab |
| Vietnam | Oral swab |

*lower mean, higher the preference rank

**Supplementary Table 7.** **Most preferred diagnostic testing method in Europe**

| Diagnostic Testing Method | Mean Values of Preference Rank* |
|---|---|
| Oral swab | 2.58 |
| Anterior-nares (AN) nasal swab | 2.49 |
| Saliva | 2.98 |
| Oropharyngeal | 3.52 |
| Nasopharyngeal | 4.61 |

| | |
|---|---|
| Deep coughing | 4.75 |
| **Top Preference by Country** | |
| Austria | Nasopharyngeal |
| Belgium | Saliva |
| Bosnia & Herzegovina | AN swab |
| Bulgaria | Saliva |
| Croatia | Saliva |
| Czech Republic | Saliva |
| Denmark | Oral swab |
| Estonia | AN swab |
| Finland | AN swab |
| France | Oral swab |
| Georgia | Oral swab |
| Germany | AN swab |
| Greece | Oral swab |
| Hungary | Oropharyngeal |
| Ireland | Oral swab |
| Italy | Saliva |
| Latvia | AN swab |
| Lithuania | AN swab |
| Luxembourg | Oral swab |
| Netherlands | Oral swab |
| Norway | Saliva |
| Poland | Oral swab |
| Portugal | Oral swab |
| Romania | Saliva |
| Russia | AN swab |
| Serbia | Saliva |
| Slovakia | Saliva |
| Slovenia | AN swab |
| Spain | Oral swab |
| Sweden | Saliva |
| Switzerland | Saliva |
| United Kingdom | AN swab |

*Lower mean, higher the preference rank

**Supplementary Table 8. Most preferred diagnostic testing method in Latin America & The Caribbean**

| Diagnostic Testing Method | Mean Values Of Preference Rank* |
|---|---|
| Saliva | 2.61 |
| Oral swab | 2.68 |
| Anterior-nares (AN) nasal swab | 3.25 |
| Oropharyngeal | 3.61 |
| Nasopharyngeal | 3.97 |
| Deep coughing | 4.68 |
| **Top Preference by Country** | |
| Argentina | AN swab |
| Barbados | Saliva |
| Bolivia | Saliva |
| Brazil | Saliva |
| Chile | Oral swab |
| Columbia | Oral swab |
| Costa Rica | Nasopharyngeal |
| Ecuador | Saliva |
| El Salvador | AN swab |
| French Guiana | AN swab |
| Guatemala | AN swab |
| Guyana | AN swab |
| Honduras | Oral swab |
| Mexico | Saliva |
| Paraguay | Saliva |
| Peru | Nasopharyngeal |
| Saint Kitts and Nevis | Oral swab |
| Trinidad and Tobago | AN swab |
| Uruguay | Oral swab |
| Venezuela | Saliva |

*Lower mean, higher the preference rank

**Supplementary Table 9. Most preferred diagnostic testing method in North America**

| Diagnostic Testing Method | Mean Values Of Preference Rank* |
|---|---|
| Oral swab | 2.28 |
| Anterior-nares (AN) nasal swab | 2.35 |
| Saliva | 2.51 |
| Oropharyngeal | 3.93 |
| Deep coughing | 4.97 |

| | |
|---|---|
| Nasopharyngeal | 2.35 |
| **Top Preference by Country** | |
| Canada | Oral swab |
| United States | Oral swab |

*Lower mean, higher the preference rank

**Supplementary Table 10. Most preferred diagnostic testing method in Africa**

| Diagnostic Testing Method | Mean Values Of Preference Rank* |
|---|---|
| Oral swab | 3.10 |
| Saliva | 3.33 |
| Anterior-nares (AN) nasal swab | 3.24 |
| Oropharyngeal | 3.37 |
| Nasopharyngeal | 3.73 |
| Deep coughing | 4.01 |
| **Top Preference by Country** | |
| Benin | Saliva |
| Botswana | Saliva |
| Burkina Faso | Nasopharyngeal |
| Cameroon | Saliva |
| Democratic Republic of the Congo | Oral swab |
| Ethiopia | AN swab |
| Gabon | Oral swab |
| Ghana | Oral swab |
| Kenya | AN swab |
| Lesotho | Saliva |
| Liberia | Saliva |
| Libya | AN swab |
| Madagascar | Oropharyngeal |
| Mali | Oropharyngeal |
| Mauritius | Nasopharyngeal |
| Morocco | AN swab |
| Mozambique | Saliva |
| Nigeria | Oral swab |
| Rwanda | Oral swab |
| Senegal | Saliva |
| Somalia | Saliva |
| South Africa | Oral swab |

| | |
|---|---|
| South Sudan | Nasopharyngeal |
| Sudan | Oral swab |
| Tanzania | Saliva |
| The Gambia | Oral swab |
| Tunisia | Nasopharyngeal |
| Uganda | Oral swab |
| Zambia | Saliva |
| Zimbabwe | Oral swab |

*Lower mean, higher the preference rank

**Supplementary Table 11. Most preferred diagnostic testing method in the Middle East**

| Diagnostic Testing Method | Mean Values Of Preference Rank* |
|---|---|
| Oral swab | 3.15 |
| Saliva | 3.36 |
| Anterior-nares (AN) nasal swab | 3.09 |
| Oropharyngeal | 3.38 |
| Nasopharyngeal | 3.58 |
| Deep coughing | 4.38 |
| **Top Preference by Country** | |
| Armenia | AN swab |
| Azerbaijan | N/A |
| Bahrain | AN swab |
| Egypt | Nasopharyngeal |
| Iran | Oral swab |
| Iraq | AN swab |
| Israel | Oral swab |
| Jordan | Oropharyngeal |
| Kuwait | Oropharyngeal |
| Lebanon (Beirut) | Saliva |
| Oman | Saliva |
| Palestine | AN swab |
| Qatar | Oral swab |
| Saudi Arabia | Oral swab |
| Syria | AN swab |
| Turkey | Saliva |
| United Arab Emirates (UAE) | AN swab |
| Yemen | Nasopharyngeal |

*Lower mean, higher the preference rank

**Supplementary Table 12.** **Most preferred diagnostic testing method in Oceania**

| Diagnostic Testing Method | Mean Values Of Preference Rank* |
|---|---|
| Saliva | 2.19 |
| Oral swab | 2.13 |
| Anterior-nares (AN) nasal swab | 3.06 |
| Oropharyngeal | 3.78 |
| Deep coughing | 5.01 |
| Nasopharyngeal | 4.92 |
| **Top Preference by Country** | |
| Australia | Oral swab |
| New Zealand | Saliva |
| Papua New Guinea | Nasopharyngeal |

*Lower mean, higher the preference rank

**Supplementary Table 13.** **Most preferred diagnostic testing method for children in Asia**

| Diagnostic Testing Method | Responses (%) |
|---|---|
| Drooling saliva into a small plastic tube (saliva) | 30.2% |
| I do not have children | 23.3% |
| Using a swab to collect a sample from about halfway up your nose (anterior-nares (AN) nasal swab) | 12.8% |
| Using a swab to collect a sample from going all the way through your nose to the back of your throat (nasopharyngeal) | 12.8% |
| Using a swab to collect a sample from just inside your mouth (oral swab) | 10.5% |
| Using a swab to collect a sample from the back of your mouth (oropharyngeal) | 8.1% |
| Deep coughing to collect fluid from in the lungs | 2.3% |
| **Top Preference by Country** | |
| Bangladesh | Nasopharyngeal |
| Bhutan | N/A |
| Cambodia | Deep Coughing |
| China | Saliva |
| India | Saliva |
| Indonesia | Saliva |
| Japan | Nasopharyngeal |
| Kazakhstan | Saliva |
| Malaysia | Saliva |
| Myanmar | Saliva |

| | |
|---|---|
| Nepal | Saliva |
| Pakistan | Nasopharyngeal |
| Philippines | Saliva |
| Singapore | Saliva |
| South Korea | Saliva |
| Sri Lanka | Saliva |
| Taiwan | Saliva |
| Tajikistan | Deep Coughing |
| Thailand | Saliva |
| Vietnam | Oral Swab |

**Supplementary Table 14. Most preferred diagnostic testing method for children in Europe**

| Diagnostic Testing Method | Responses (%) |
|---|---|
| I do not have children | 40.7% |
| Using a swab to collect a sample from about halfway up your nose (anterior-nares (AN) nasal swab) | 20.4% |
| Drooling saliva into a small plastic tube (saliva) | 16.7% |
| Using a swab to collect a sample from just inside your mouth (oral swab) | 16.7% |
| Using a swab to collect a sample from going all the way through your nose to the back of your throat (nasopharyngeal) | 3.7% |
| Using a swab to collect a sample from the back of your mouth (oropharyngeal) | 0% |
| Deep coughing to collect fluid from in the lungs | 0% |
| **Top Preference by Country** | |
| Austria | Saliva |
| Belgium | Oral swab |
| Bosnia & Herzegovina | Nasopharyngeal |
| Bulgaria | Deep Coughing |
| Croatia | Saliva |
| Czech Republic | Saliva |
| Denmark | Saliva |
| Estonia | Deep Coughing |
| Finland | AN swab |
| France | Oral Swab |
| Georgia | Oral Swab |
| Germany | AN swab |
| Greece | AN swab |
| Hungary | Deep Coughing |
| Ireland | Saliva |

| | |
|---|---|
| Italy | Saliva |
| Latvia | AN swab |
| Lithuania | AN swab |
| Luxembourg | N/A |
| Netherlands | AN swab |
| Norway | Deep Coughing |
| Poland | Deep Coughing |
| Portugal | Oral Swab |
| Romania | Saliva |
| Russia | Oral Swab |
| Serbia | N/A |
| Slovakia | Saliva |
| Slovenia | AN swab |
| Spain | Saliva |
| Sweden | Nasopharyngeal |
| Switzerland | Saliva |
| United Kingdom | AN swab |

**Supplementary Table 15. Most preferred diagnostic testing method for children in Latin America & The Caribbean**

| Diagnostic Testing Method | Responses (%) |
|---|---|
| I do not have children | 37.1% |
| Drooling saliva into a small plastic tube (saliva) | 21.3% |
| Using a swab to collect a sample from just inside your mouth (oral swab) | 20.2% |
| Using a swab to collect a sample from going all the way through your nose to the back of your throat (nasopharyngeal) | 13.5% |
| Using a swab to collect a sample from about halfway up your nose (anterior-nares (AN) nasal swab) | 6.7% |
| Using a swab to collect a sample from the back of your mouth (oropharyngeal) | 1.1% |
| Deep coughing to collect fluid from in the lungs | 0% |
| **Top Preference by Country** | |
| Argentina | I do not have children |
| Barbados | AN swab |
| Bolivia | AN swab |
| Brazil | Saliva |
| Chile | Saliva |
| Columbia | Saliva |
| Costa Rica | Nasopharyngeal |

| | |
|---|---|
| Ecuador | Nasopharyngeal |
| El Salvador | Oropharyngeal |
| French Guiana | Oral Swab |
| Guatemala | AN swab |
| Guyana | Oral Swab |
| Honduras | Saliva |
| Mexico | Oral Swab |
| Paraguay | Saliva |
| Peru | Saliva |
| Saint Kitts and Nevis | Oral Swab |
| Trinidad and Tobago | Nasopharyngeal |
| Uruguay | Saliva |
| Venezuela | Nasopharyngeal |

**Supplementary Table 16. Most preferred diagnostic testing method for children in North America**

| Diagnostic Testing Method | Responses (%) |
|---|---|
| I do not have children | 50.5% |
| Drooling saliva into a small plastic tube (saliva) | 19.3% |
| Using a swab to collect a sample from just inside your mouth (oral swab) | 15.8% |
| Using a swab to collect a sample from about halfway up your nose (anterior-nares (AN) nasal swab) | 12.1% |
| Using a swab to collect a sample from going all the way through your nose to the back of your throat (nasopharyngeal) | 1.0% |
| Using a swab to collect a sample from the back of your mouth (oropharyngeal) | 1.0% |
| Deep coughing to collect fluid from in the lungs | 0.21% |
| **Top Preference by Country** | |
| Canada | Oral Swab |
| United States | Saliva |

**Supplementary Table 17. Most preferred diagnostic testing method for children in Africa**

| Diagnostic Testing Method | Responses (%) |
|---|---|
| Drooling saliva into a small plastic tube (saliva) | 26.7% |
| Using a swab to collect a sample from just inside your mouth (oral swab) | 21.7% |
| I do not have children | 20.8% |
| Using a swab to collect a sample from the back of your mouth (oropharyngeal) | 13.3% |
| Using a swab to collect a sample from about halfway up your nose (anterior-nares (AN) nasal swab) | 10.8% |
| Using a swab to collect a sample from going all the way through your nose to the back of your throat (nasopharyngeal) | 3.3% |

| | |
|---|---|
| Deep coughing to collect fluid from in the lungs | 3.3% |
| **Top Preference by Country** | |
| Benin | Saliva |
| Botswana | Saliva |
| Burkina Faso | Oropharyngeal |
| Cameroon | AN swab |
| Democratic Republic of the Congo | AN swab |
| Ethiopia | Saliva |
| Gabon | Oral swab |
| Ghana | Oral swab |
| Kenya | Saliva |
| Lesotho | Saliva |
| Liberia | Saliva |
| Libya | AN swab |
| Madagascar | Saliva |
| Mali | Oral swab |
| Mauritius | Nasopharyngeal |
| Morocco | Saliva |
| Mozambique | Saliva |
| Nigeria | Saliva |
| Rwanda | Oral swab |
| Senegal | Saliva |
| Somalia | AN swab |
| South Africa | Oral swab |
| South Sudan | Oral swab |
| Sudan | Oral swab |
| Tanzania | Saliva |
| The Gambia | Oral swab |
| Tunisia | Oral swab |
| Uganda | Saliva |
| Zambia | Saliva |
| Zimbabwe | Deep coughing |

**Supplementary Table 18. Most preferred diagnostic testing method for children in the Middle East**

| Diagnostic Testing Method | Responses (%) |
|---|---|
| Drooling saliva into a small plastic tube (saliva) | 19.3% |
| Using a swab to collect a sample from just inside your mouth (oral swab) | 19.3% |

| | |
|---|---|
| I do not have children | 30.7% |
| Using a swab to collect a sample from the back of your mouth (oropharyngeal) | 7.3% |
| Using a swab to collect a sample from about halfway up your nose (anterior-nares (AN) nasal swab) | 10.0% |
| Using a swab to collect a sample from going all the way through your nose to the back of your throat (nasopharyngeal) | 13.3% |
| Deep coughing to collect fluid from in the lungs | 0% |
| **Top Preference by Country** | |
| Armenia | AN swab |
| Azerbaijan | N/A |
| Bahrain | AN swab |
| Egypt | Oropharyngeal |
| Iran | Saliva |
| Iraq | AN swab |
| Israel | Oral swab |
| Jordan | Saliva |
| Kuwait | Oral swab |
| Lebanon (Beirut) | Oral swab |
| Oman | Saliva |
| Palestine | Oral swab |
| Qatar | Saliva |
| Saudi Arabia | Saliva |
| Syria | Deep coughing |
| Turkey | Saliva |
| United Arab Emirates (UAE) | Saliva |
| Yemen | Nasopharyngeal |

**Supplementary Table 19. Most preferred diagnostic testing method for children in Oceania**

| Diagnostic Testing Method | Responses (%) |
|---|---|
| Drooling saliva into a small plastic tube (saliva) | 21.0% |
| Using a swab to collect a sample from just inside your mouth (oral swab) | 15.2% |
| Using a swab to collect a sample from about halfway up your nose (anterior-nares (AN) nasal swab) | 7.6% |
| Using a swab to collect a sample from going all the way through your nose to the back of your throat (nasopharyngeal) | 2.0% |
| Using a swab to collect a sample from the back of your mouth (oropharyngeal) | 0.9% |
| Deep coughing to collect fluid from in the lungs | 0% |
| I do not have children | 53.3% |
| **Top Preference by Country** | |

| | |
|---|---|
| Australia | Saliva |
| New Zealand | Saliva |
| Papua New Guinea | Nasopharyngeal |
