## Appendix for "Diagnostic testing preferences can help inform future public health response efforts: global insights from an international survey"

### APPENDIX:
### Survey Questions

#### Diagnostic testing preferences can help inform future public health response efforts: global insights from an international survey

Leah Salzano[1]*, Nithya Narayanan[1]*, Emily R. Tobik[1]*, Sumaira Akbarzada[1], Yanjun Wu[1], Sarah Megiel[1], Brittany Choate[1,2], Anne L. Wyllie[1]

[1]Department of Epidemiology of Microbial Diseases, Yale School of Public Health, New Haven, CT 06510, USA;
[2]Department of Health Policy and Management, Yale School of Public Health, New Haven, CT 06510, USA;
[3]SalivaDirect, Inc., New Haven, CT 06510, USA.

*These authors contributed equally

Corresponding Author:
Anne Wyllie
Yale School of Public Health
LEPH 823
New Haven
CT 06510
203-781-6948


### Questionnaire

What is your age?

○ Under 18 (1)

○ 18 - 24 (2)

○ 25 - 34 (3)

○ 35 - 44 (4)

○ 45 - 54 (5)

○ 55 - 64 (6)

○ 65+ (7)

*Skip To: End of Survey If What is your age? = Under 18*

What is your gender?

- ○ Male (1)
- ○ Female (2)
- ○ Non-binary (3)
- ○ Other (4)
- ○ Prefer not to respond (5)

What is your degree of educational attainment?

- ○ No schooling completed (9)
- ○ Some primary or elementary school (11)
- ○ Some secondary or high school (1)
- ○ High school graduate or equivalent (2)
- ○ Some college, no degree (3)
- ○ Trade school (4)
- ○ Associate degree (5)
- ○ Bachelor's degree (6)
- ○ Graduate degree (7)
- ○ Prefer not to say (8)

What is your race/ethnicity? (select all that apply)

- ☐ African (11)
- ☐ Black/African American (17)
- ☐ Caribbean (12)
- ☐ East Asian (14)
- ☐ Latino/Hispanic (9)
- ☐ Middle Eastern (10)
- ☐ South Asian (13)
- ☐ White/Caucasian (8)
- ☐ Other (6)
- ☐ Prefer not to say (7)

What country and city do you currently live in? (Please write out the full country and city name)

________________________________________________________________

If you are living abroad, what is your country of origin?

________________________________________________________________

What is your employment status?

- ○ Employed, full-time (4)

- Employed, part-time (1)
- Homemaker or caregiver (11)
- Self-employed (10)
- Not employed, looking for work (5)
- Not employed, not looking for work (6)
- Student (7)
- Retired (8)
- Disabled, not able to work (9)

*Display This Question:*

*If What is your employment status? = Employed, full-time*

*Or What is your employment status? = Employed, part-time*

*Or What is your employment status? = Self-employed*

*Or What is your employment status? = Not employed, looking for work*

Pick which best describes your occupational group:

- Arts (1)
- Agriculture (4)
- Construction (5)
- Clergy (6)
- Education (12)
- Food Preparation / Service (7)
- Healthcare Worker (8)

- ◯ Law enforcement (14)
- ◯ Science / Research (9)
- ◯ Financial Services / Accounting (11)
- ◯ Management (19)
- ◯ Manufacturing / Industrial Work (10)
- ◯ Military (13)
- ◯ Legal (15)
- ◯ Customer service (16)
- ◯ Retail / Sales (17)
- ◯ Administration (18)
- ◯ Other (22)

If you had to go for a COVID-19 test tomorrow, what type of test would you prefer/seek out?

________________________________________________________________

Do you have a preferred brand of COVID-19 test? (e.g. Abbott, LabCorp, Roche, Quidel, etc.)

________________________________________________________________

What is important to you in a COVID-19 test?

________________________________________________________________

Please refer to these images [available upon request from the authors], as needed, to help inform your responses to the questions below.

Which of the following COVID testing options have you heard of? (select all that apply)

- ☐ Using a swab to collect a sample from about halfway up your nose (anterior-nares (AN) nasal swab, graphic 1) (1)
- ☐ Using a swab to collect a sample from going all the way through your nose to the back of your throat (nasopharyngeal swab, graphic 2) (2)
- ☐ Using a swab to collect a sample from just inside your mouth (oral swab, graphic 3) (4)
- ☐ Using a swab to collect a sample from the back of your mouth (oropharyngeal swab, graphic 4) (3)
- ☐ Drooling saliva into a small plastic tube (saliva testing, graphic 5) (5)
- ☐ Deep coughing to collect fluid from in the lungs (6)

Which of the following COVID testing options are offered in your area? (select all that apply)

| | PCR (1) | Rapid Antigen (2) |
|---|---|---|
| Nasal swab (4) | ☐ | ☐ |
| Nasopharyngeal swab (1) | ☐ | ☐ |
| Oral swab (5) | ☐ | ☐ |
| Oropharyngeal swab (2) | ☐ | ☐ |
| Saliva test (6) | ☐ | ☐ |

Which of the following COVID testing options have you used? (select all that apply)

| | PCR (1) | Rapid Antigen (2) |
|---|---|---|
| Nasal swab (4) | ☐ | ☐ |
| Nasopharyngeal swab (1) | ☐ | ☐ |
| Oral swab (5) | ☐ | ☐ |
| Oropharyngeal swab (2) | ☐ | ☐ |
| Saliva test (6) | ☐ | ☐ |

Which of the following COVID testing options do you trust to provide accurate results? (select all that apply)

☐ PCR (1)

☐ Rapid antigen test (2)

☐ Neither (3)

☐ Not sure (4)

Which of the following sample types would you trust to provide accurate results? (select all that apply)

☐ Nasal swab (1)

☐ Nasopharyngeal swab (2)

☐ Oral swab (3)

☐ Oropharyngeal swab (4)

☐ Saliva test (5)

Assuming you would receive equally accurate results from all options, which of the following COVID testing options do you prefer? Please rank the options from most preferred (1) to least preferred (6).

_______ Using a swab to collect a sample from about halfway up your nose (anterior-nares (AN) nasal swab) (1)

_______ Using a swab to collect a sample from going all the way through your nose to the back of your throat (nasopharyngeal) (2)

_______ Using a swab to collect a sample from the back of your mouth (oropharyngeal) (3)

_______ Using a swab to collect a sample from just inside your mouth (oral swab) (4)

_______ Drooling saliva into a small plastic tube (saliva testing) (5)

_______ Deep coughing to collect fluid from in the lungs (6)

If you had a negative test but started to develop additional symptoms, how likely would you be to go for a follow-up test if it required the following sample? Please rank the options from most likely (1) to least likely (6).

_______ A swab to collect a sample from about halfway up your nose (anterior-nares (AN) nasal swab) (1)

_______ A swab to collect a sample from going all the way through your nose to the back of your throat (nasopharyngeal) (2)

_______ A swab to collect a sample from the back of your mouth (oropharyngeal) (3)

_______ A swab to collect a sample from just inside your mouth (oral swab) (4)

_______ Drooling saliva into a small plastic tube (saliva testing) (5)

_______ Deep coughing to collect fluid from in the lungs (6)

Do you prefer...

○ To have a sample collected for a COVID test by a medical professional (1)

○ To self-collect your own sample for a COVID test (2)

○ No preference (3)

If you have children, which of the following COVID testing options would you MOST prefer for your child?

- ◯ Using a swab to collect a sample from about halfway up your nose (anterior-nares (AN) nasal swab) (1)
- ◯ Using a swab to collect a sample from going all the way through your nose to the back of your throat (nasopharyngeal) (2)
- ◯ Using a swab to collect a sample from the back of your mouth (oropharyngeal) (3)
- ◯ Using a swab to collect a sample from just inside your mouth (oral swab) (4)
- ◯ Drooling saliva into a small plastic tube (saliva testing) (5)
- ◯ Deep coughing to collect fluid from in the lungs (6)
- ◯ I do not have children (7)

The survey will be submitted on the next page. Please review your answers above before submitting.
